# Drug-combination wide association studies of cancer

**DOI:** 10.1101/2022.09.29.22280479

**Authors:** Panagiotis Nikolaos Lalagkas, Rachel Dania Melamed

## Abstract

**Background:** Combinations of common drugs may, when taken together, have unexpected effects on cancer. It is not feasible to test for all combination drug effects in clinical trials, but in the real world, drugs are frequently taken in combination. Then, there may be undiscovered effects protecting users from cancer–or increasing their risk. By analyzing massive health claims data containing numerous people exposed to drug combinations, we have an opportunity to evaluate the association of drug combinations with cancer risk. Discovering these effects can not only contribute to prevention of cancer, but also suggest new uses for combinations to prevent or treat cancer.

**Method:** Our approach emulates a randomized trial where one arm would have been assigned to take a particular drug alone, while the other arm takes it together with a second drug. Because discovery of associations from observational data is prone to spurious results due to confounding, we develop strategies to distinguish confounding from biomedically relevant findings.

**Results:** This tactic allows us to systematically assess effects across over 9,000 drug combinations, on all common cancers. Through multiple sensitivity analyses we identify a robustly supported beneficial drug combination that may synergistically impact lipid levels to reduce risk of cancer.

**Conclusions:** This study demonstrates the importance of considering confounding factors in drug-wide studies. But, we also show that our method is able to uncover associations with robust support.

**Impact:** Searching for combinations of factors impacting cancer is crucial, but these effects can only be systematically discovered through observational data analysis.

## Introduction

Late onset chronic diseases are responsible for a bulk of deaths, with cancer ranking as the second leading cause of death^1^. Despite intensive research, we have not identified effective ways of preventing most cases, and new treatments are urgently needed. To address this problem, one approach is to investigate what common exposures could be associated with disease risk. Among people over 40 years old, one third use two or more drugs, and 20% use more than five medications^2^. We propose that combinations of common medications could influence cancer risk.

These effects are unlikely to have been uncovered in randomized trials, partly due to the short follow-up period of most trials^3,4^. However, some drug combinations have been found to impact cancers. While estrogen replacement as a treatment for menopause is not linked to adverse cancer outcomes, combining estrogen with progesterone appeared to increase risk of breast cancer over a three year follow-up period^5^. This effect may be explained biologically: estrogen can promote proliferation, while progesterone enhances angiogenesis in the proliferating tissue^6^. In addition to such deleterious effects, drug combinations can cause hidden benefits. Statins, prescribed for high cholesterol, have been linked to reduced incidence of multiple cancer types in epidemiological studies, and they are in trials for cancer therapy. Trials have investigated anti-cancer effects of statins combined with celecoxib (anti-inflammatory) and metformin (a diabetic drug)^7^. These examples describe combinations of common drugs positively and negatively impacting cancer.

As many thousands of common drug combinations could be tested for any of a dozen types of common cancer, these effects are unlikely to be discovered through experimental trials. Outside of the setting of clinical trials, one approach to discovering such drug effects is analysis of observational data. Using such health data, we can follow health outcomes in people exposed to the drug combination. Although reusing existing data is fast, cheap, and allows systematic discovery, new methods are needed to find drug combination effects. Unlike experimental data, observational data is subject to confounding: people are prescribed drugs based on their health, and some health conditions incur increased risk of cancer. For instance, smoking increases lung cancer risk, meaning that people who take smoking cessation drugs are likely at increased risk of lung cancer. Without accounting for such variation in health, the drugs and drug combinations most associated with cancer would be dominated by such spurious associations.

The typical approach to discovering a biomedically relevant association from observational data is to attempt to emulate a randomized trial that could have investigated the same question^8^. Two study cohorts, consisting of users of a drug, or some control group are “enrolled”, and then the analysis compares their outcomes, adjusting for confounders of the association. While we do not know of other drug combination-wide studies, we have reviewed previous drug-wide association studies of cancer (single drug, not combinations)^9–12^. Because of their systematic nature, these studies make some simplifying assumptions. Particularly, they often adjust for a limited set of pre-defined confounders. It is not clear if these measures adequately adjust for confounding. To attempt to detect and resolve bias due to remaining confounding, a tactic called “empirical calibration” compares the effect of a drug on outcomes of interest against its association with selected “negative control” outcomes^13,14^. For instance, the authors expect most common drugs do not affect risk of ingrown nails, so any association between medications and ingrown nail can be attributed to bias. This method uses effect estimates across negative control outcomes to approximate, and thus remove, the bias. This procedure requires the assumption that all effect estimates are centered around a single overall bias. One weakness of this assumption is that bias is often specific to the particular pair of exposure and outcome. For instance, people taking smoking cessation drugs are at risk of lung cancer, but not breast cancer. As well, randomly selected negative controls may not suffer the same bias; we expect no confounding effect associating smoking cessation agents with ingrown nail.

Therefore, here we develop new methods for *drug combination-wide association studies*. Our aim is a systematic discovery of drug combinations that, like other drug-wide methods, allows unprejudiced investigation of the effect of any observable drug combination. In contrast to other drug-wide methods, we make fewer assumptions about the nature of confounding: 1) we consider that any event in medical history could be a possible confounding factor; 2) we do not assume that all associations are subject to the same type of confounding. We accept that due to the nature of observational data, our results must be replicated and verified by future experimental studies. But, we make extensive efforts to identify remaining confounding, and we use a number of sensitivity analysis to pinpoint associations that are most robustly supported. In addition to these novel advances, this work is the first to develop a new method to systematically discover interaction effects of drugs on disease. We apply these methods to one of the largest available compilations of health care data, the Truven MarketScan data set containing over 100 million individuals. We expect both our methods and the identified drug combination associations can provide advances toward clinically relevant insights from observational data.

## Results

### Observational data to emulate a randomized trial of drug combinations

To develop our approach, we first consider a hypothetical randomized trial that would test the association of any arbitrary single drug with a common cancer outcome. A hypothetical experimental design could enroll people never exposed to the drug of interest (such as a statin), and then randomize some to take statin, and others to remain unexposed, and then follow time to cancer (*Figure 1A*). Previous drug-wide studies of cancer using observational data have emulated this type of experimental study using either cohort study approach or a case-control approach^9–12^. Typically, they select an index date to assess exposure and create the exposed and unexposed cohorts. Then, time after the index date is considered time at-risk for the outcome. This design allows us to assess whether a particular drug is associated with cancer, and the same procedure is repeated for every common drug. These studies typically adjust for a handful of possible confounding factors (such as age, and Charlson comorbidity index). For example, in the study of Patel, et al., the authors followed people in a prescription database, assessing exposure to drug as a time-varying covariate, and adjusting for age, sex, and presence of any other prescriptions^9^. Støer, et. al., took a case-control approach, matching cancer cases to non-cancers, and assessing presence of exposure based on prescription history up to one year before cancer^11^. These designs all have the same basic goal of estimating the result of the trial described in *Figure 1A*.

**Figure 1.**
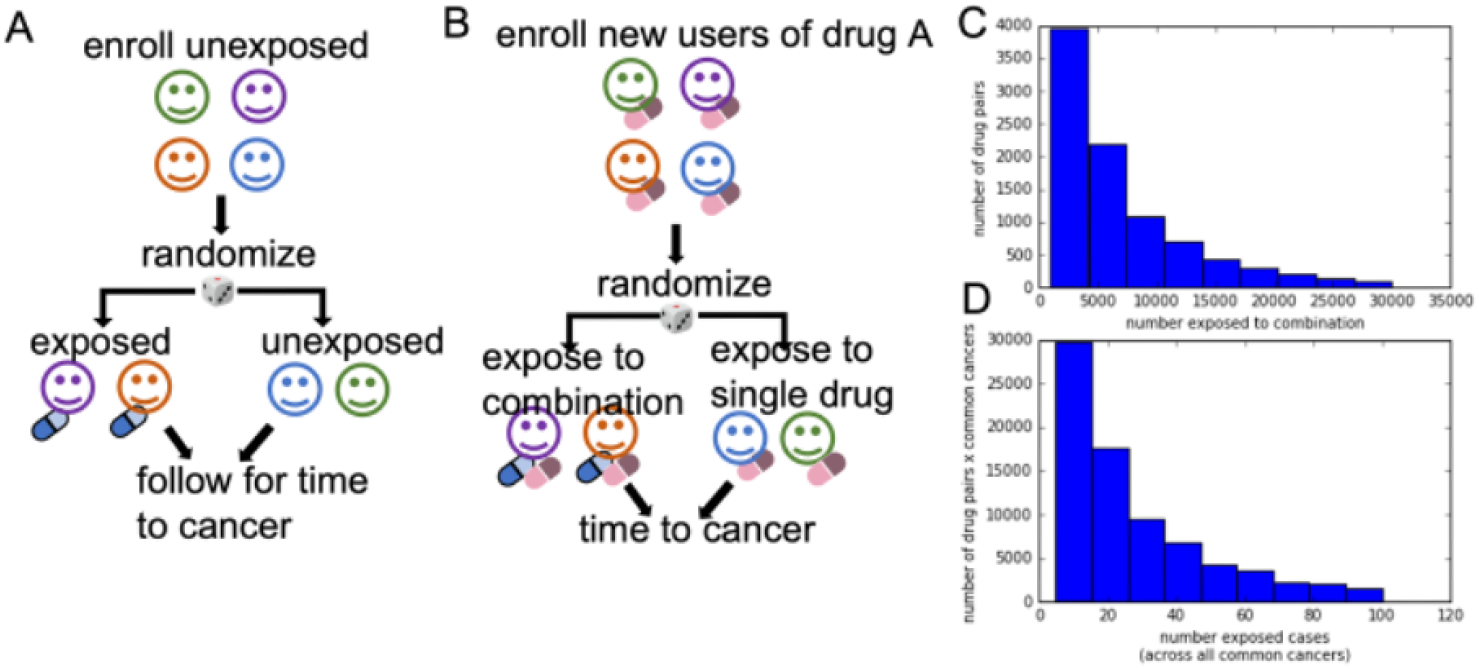
**A**. Hypothetical randomized design for assessing drug effect, can be applied to estimate effects of any common drug**. B** Hypothetical randomized trial of combinations of common drugs**. C**. Total numbers of people exposed to a drug pair, across 9,502 drug pairs. **D**. Number of cancer cases per drug pair, aggregated across all common cancers

Building on these works, we design a new hypothetical randomized trial to estimate the effect of a combination of an arbitrary pair of drugs A and B. First, we would enroll new users of some drug A who have never before taken either drug A or B, and with no history of cancer. Then, the hypothetical trial would randomly assign some people to additionally take drug B (*Figure 1B*). Finally, we would follow these two groups to observe incidence of cancer. This approach is most similar to the method of Patel, et al., but we make extensive effort to adjust for confounding. Analyses of the results could follow all people who were initially randomized, called an *intention-to-treat* analysis. An alternative analysis could censor follow-up time for those who discontinued use of drug A, known as a *per-protocol* analysis.

To emulate such a trial, we use a set of health claims data from Truven MarketScan (now IBM), containing 150 million drug users. We obtain coded diagnoses, prescriptions, and procedures, alongside basic demographic characteristics, over a twelve year span from 2003 to 2015. We observe 9,502 drug combinations with a median of 4,979 combination users per drug pair (*Figure 1C*), and with a median of 20 exposed people who eventually develop a cancer (*Figure 1D*). To summarize, we perform this emulation of a randomized trial for 9,502 pairs of a drug A and B. For the rest of this manuscript, we refer to the first drug as drug A, and the second drug as drug B. In theory, we might expect the same true effect if the order of the drugs were reversed, but in practice sample size limitations mean we usually do not observe both orders.

### Approach to emulating the randomized trial

We divide our main design into three steps, analogous to steps of a randomized trial: 1) enrollment (creation of our cohorts); 2) randomization (adjusting for time-varying confounding); and 3) analysis of outcomes and estimation of effects.

In the enrollment step, as mentioned, we compile all new users of drug A, who have never taken drug B, or had the outcomes of interest. Then, we follow this cohort over time, seeing whether those who additionally take drug B alongside A (treated group) have different cancer outcomes than those who take just drug A (comparator group). While in the randomized setting, some subset would be immediately assigned to take the combination of drugs A and B, in the observational setting, some people will start drug B immediately, while others might start drug B later. While we could only compare those who start drug B immediately against those who continue on drug A alone, this choice would drastically reduce our data size, making it impossible to assess the effect of most drug combinations. In order to capture all person-time under joint exposure of the drug combination, we follow the established approach of emulating a *sequence of* randomized trials^15,16^. This tactic divides follow-up time after initiation of drug A for each person into arbitrary but reasonable windows (we choose 3 or 6 month windows). Each time window defines a trial in the sequence of trials: a person who has not initiated drug B yet at follow-up time window *w* remains eligible to “enroll” in a trial that starts at that time window (*Figure 2A*). Therefore, we are able to include all joint users of drugs A and B in one of these trials. For instance, person i (*Figure 2A,B*) who started the drug combination in window 2 is in the treated group for trial 2, but is in the comparator group for trial 1. This person is not eligible to join trials starting after trial 2, because at that point they have a history of taking the drug combination (an exclusion criterion). We can easily account for this replicated data by indicating the repeated observations in our statistical analysis (see Methods).

**Figure 2.**
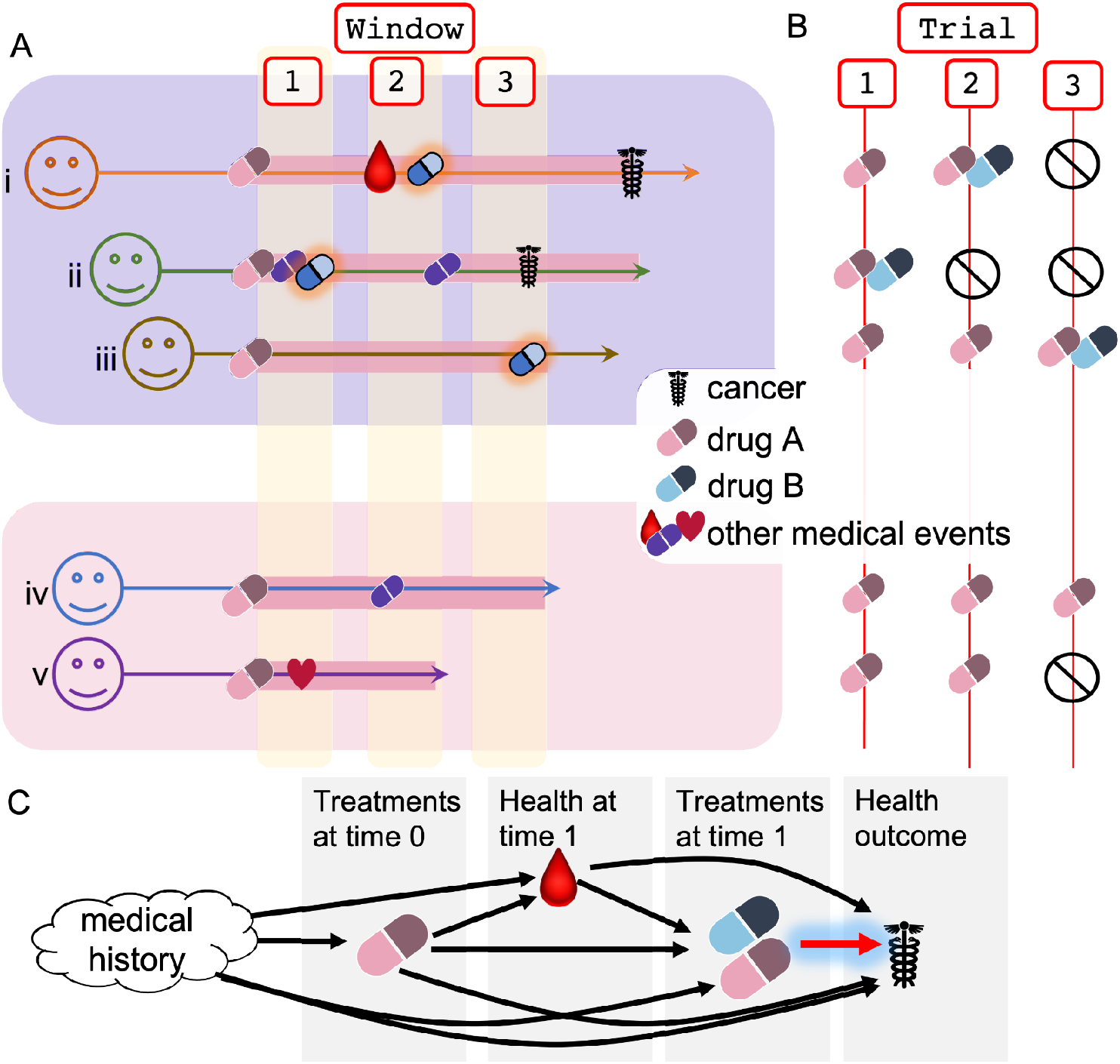
Enrollment for trial and addressing confounding. **A**. Medical history for five people exposed to drug A (pink pill) is represented with time along the horizontal axis. Follow-up time is divided into 3 windows. The top three people are eventually exposed to the combination of drugs A and B (blue pill), while the bottom two people remain in the comparator group (drug A alone). **B**. Representation of the expansion of person-time for these five people into a sequence of randomized trials. Person 1 is unexposed at time window 1 (trial 1), and thus is in the comparator group, but still eligible to participate in trial 2. At trial 2 they receive the combination, and thus are assigned to the joint-exposure group in trial 2, and they are not eligible to participate in trial 3. **C**. Each arrow represents influence or possible influence of one event in health history on another event. We assume that all items in health history can influence what happens later in time. The main edge of interest is the one highlighted in red, and all other edges represent nuisance associations. To remove confounding edges, we model probability of treatment at any time point *t*: p(exposed_t_ | health history before *t*)

In the randomization step, we emulate randomization by adjusting for time varying confounding. It is crucial to not only adjust for disparities between the treated and comparator groups at baseline initiation of drug A. This is because changes in health over time can influence which people are prescribed drug B (*Figure 2C*). For instance, onset of menopause (“Health at time 1” in *Figure 2C*) may lead to hot flashes, which could be treated with a drug like gabapentin. Menopause also increases risk of breast cancer. Therefore, if we do not adjust for time-varying confounding we would find a spurious association between gabapentin and breast cancer. Previous drug-wide methods that analyze use of single drugs alone have not required time-varying analysis, and these methods have adjusted only for selected confounders. In order to perform a drug-wide scan, we must consider that many confounders, including unexpected confounders, could affect prescription. Therefore, we consider that all medical history before time window *w* could include possible confounders. Because of the time-specific nature of medical data, colliders^14^ are unlikely: this would require both the treatment and the exposure to affect some variable that occurs *before* either of them. While errors in the medical record induce some cases of reverse causality, below we describe measures taken to guard against this issue.

To model confounding, we calculate probability of initiating drug B, based on both time window-specific and overall medical history. Confounding also can influence whether someone discontinues drug A, an issue that also arises in analysis of the data from a randomized trial. In a per-protocol analysis of randomized trials, person-time is censored after discontinuation of drug A. In order to best capture the effect of exposure to drug combinations, we also censor person-time after discontinuation of drug A. To account for confounding impacting censoring, we also model probability of discontinuation in the same way as we model initiation of drug B. Finally, we obtain time to cancer outcome using a survival analysis. The probabilities calculated in the randomization step are used as weights to remove the confounding effects associating drug and outcome in a marginal structural model^8^. In this manner, we can estimate the association between each drug combination and each common cancer, for those combinations where enough cases are observed.

### Drug-wide studies may be influenced by a wide range of confounders

Our analysis yields estimates for 9,502 drug combinations, for a median of 22 cancers where more than 100 people in the cohort study were diagnosed with that type of cancer. As mentioned, previous drug-wide studies typically account for only a handful of confounders. We investigate the possible effects of considering a narrow range of confounders. In *Figure 3A*, we compare the effect estimates that consider no confounders (unweighted); those derived from cohort studies that only adjust for age, sex, year, and Charlson comorbidity index (minimal confounding control); and our method of including a wide range of possible confounders (weighted estimates). We find that globally, after accounting for confounding, effect estimates are greatly reduced. Because this is a general effect, across many drug combinations and cancer outcomes, we conclude that this is likely to be due to a number of assorted confounding effects, rather than due to widespread effects of drug combinations of cancer. This suggests that other drug-wide studies would benefit from considering a wide range of confounders. However, the results also suggest that there is likely substantial remaining confounding: the observed linear relationship in *Figure 3B* suggests that globally, the strongest associations between treatment and outcome before adjusting for confounding are still the most associated after adjusting for confounding. While this observation shows that we need further work to avoid this bias (discussed below), this finding also underlines the importance of our drug-wide analysis. That is, the ability to assess global confounding is a key advantage of taking a systematic drug (combination)-wide approach to discovery.

**Figure 3.**
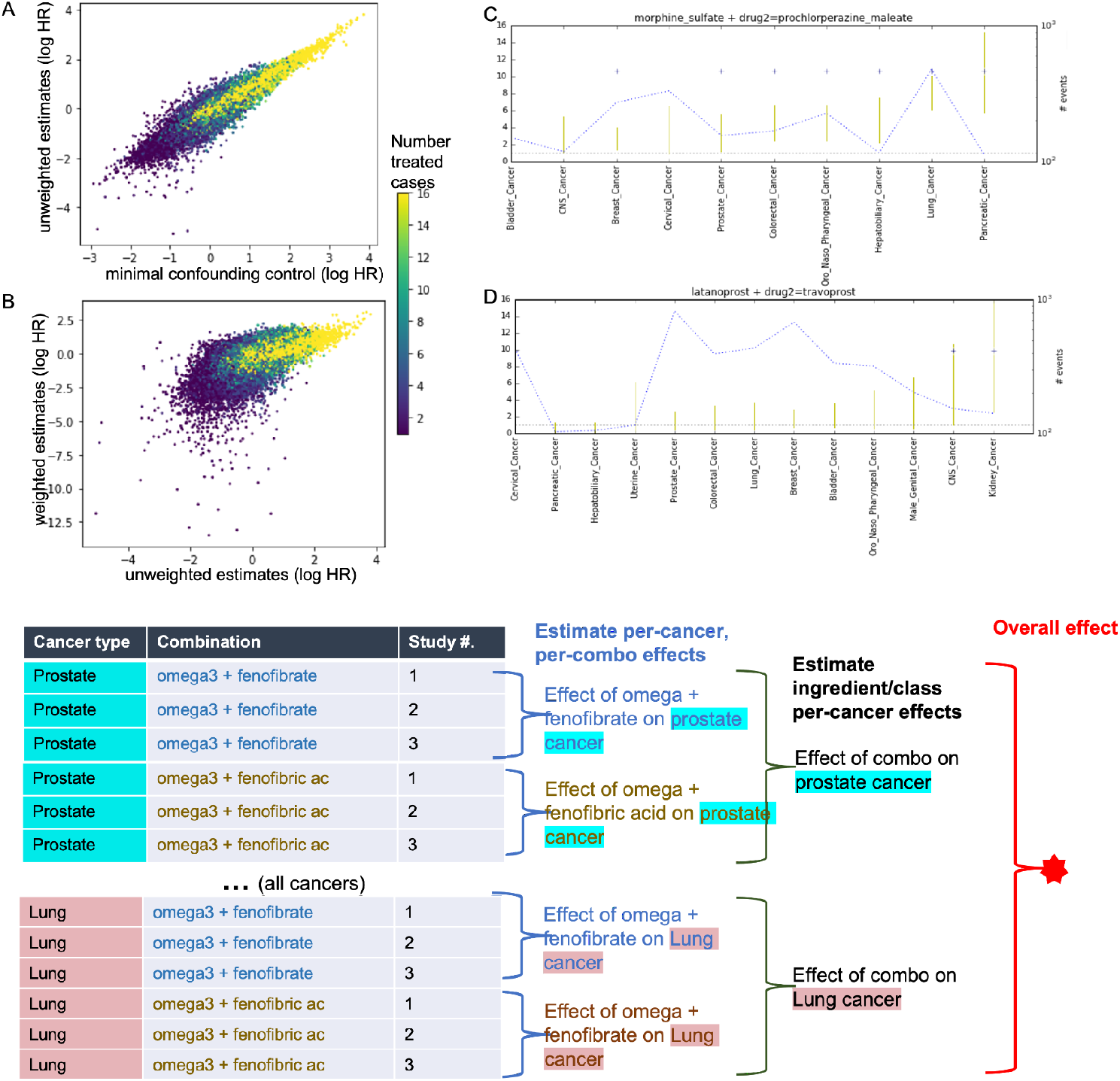
Each point is the effect estimate of one drug combination on one outcome, across 200,000 cohort studies. **A.** The unweighted and minimally adjusted estimates are largely similar to each other. **B**. **G**lobal reduction in effect estimates with weighting, due to control for confounding. By highlighting the number of treated cases, we can see that extreme “protective” (negative log hazard ratio) effects are associated with a low number of exposed cases, and likely largely spurious. **C**. Likely confounding driving drug combinations associated with cancer. Each yellow line represents the confidence interval of effect of one drug combination on cancer (hazard ratio scale, + symbols denote significance). The dashed blue line indicates number of incident cases (events) of each cancer type. **E**. A hierarchical model for identifying a combination of drug A (here omega 3 acids (fish oil)) combined with any drug B containing fenofibric acid as an active ingredient.

### Initial results of the drug-combination wide scan and remaining confounding

Across around 200,000 resulting effect estimates, we investigate selected examples to assess the nature of the remaining confounding. Among drugs most associated with increased risk of cancer, we find two obvious patterns. First, drugs used in the treatment of cancer appear associated with rates of multiple types of cancer. For example, fentanyl, morphine, and other pain treatments, as well as anti-nausea medications such as prochlorzerpine or ondansetron appear co-prescribed in people who develop cancer. This is likely due to incompleteness of the medical record: these people are overwhelmingly likely to be under treatment for cancer at the time of prescription. If the medical record had accurately recorded their cancer diagnosis, then they would not have been included in our study. Then, this is a case of *reverse causality*, where the medications are a result of the diagnosis, rather than the other way around. In selected cases of suspected reverse causality we find that the drug combination is associated with heightened risk of multiple common cancers (*Figure 3C*). This is because of shared clinical management of these diseases; many cancers are treated with surgery, chemotherapy, and drugs to manage the symptoms and complications of these procedures. We later present a simple heuristic to sort these cases.

The second pattern includes likely cases of confounding, which is usually specific to one type of cancer. For instance, complications of kidney disease include glaucoma and kidney cancer. We find that people taking a combination of two glaucoma treatments (latanaprost and travaprost), as compared to those who take only latanoprost, are at higher risk of kidney cancer (*Figure 3D*). This is overwhelmingly likely to be due to the confounding effect of kidney disease on risk of both kidney cancer, and glaucoma (and thus these drugs). Note that this combination is specifically associated with kidney cancer, which is logical due to the specific confounding effect of kidney disease. Such examples indicate remaining confounding, a separate issue from reverse causality. Again, our systematic exposure-wide assessment enables identification of this issue.

Because the presence of confounding in observational studies is well known, approaches like the Hill criteria seek to distinguish epidemiologic signals with strongest evidence of biomedically relevant effects^18^. We implement these concepts in the medication-wide setting, focusing on two criteria: consistency and specificity.

#### Consistency

The consistency criterion posits that a true effect should result in consistently observed signals, such as in repeated studies or in similar contexts. Some previous findings of drugs impacting cancer, like statins, include medications that consistently impact multiple cancer types^19^. Our examples of confounding (rather than reverse causality), on the other hand, are specific to a single cancer. Therefore, our current analysis seeks drug effects that are consistent across multiple common cancer types. As well, we consider consistency among drugs sharing an active ingredient as another form of positive signal.

Taking inspiration from the consistency heuristic, we implement an analytical approach to integrate the results of multiple cohort studies and distinguish the strongest signals from confounding. Because of the large size of the data when expanded into a sequence of windows of person-time, we split up very large cohort studies (more than about 3 million person-time points) into *replicate* cohort studies containing disjoint study populations. We integrate these multiple effect estimates in a new Bayesian hierarchical model, to estimate the per-combination effects on any cancer. The first level of the hierarchy assumes that the replicate cohort study effects of the drug combination on each common cancer are centered around the true per-cancer effect of that combination. Then, at the next level, our model specifies that effects of the combination on a particular cancer type are centered around the overall pan-cancer effect of the drug combination. In a slight variation on this model (*Figure 3E*), we estimate the effects for a combination of drug A and a drug active ingredient B, by combining the effects of all drugs that share this active ingredient. This model only differs from the previous model in that it has one extra layer in the hierarchy: we assume that the per-cancer effects for the each drug with the ingredient are drawn from the overall effect of that ingredient class on that type of cancer. The effect estimates (and their uncertainties) yielded by the Cox regression comprise the data input to the model. After fitting the model to the effect estimates, we obtain the posterior distribution of the overall effect of the combination on cancer (red star in *Figure 3E*). We classify protective drugs as those where the upper 1% posterior interval is less than zero, and predisposing drugs as those where the bottom 1% is greater than zero.

#### Specificity

Second, another criterion examines whether the associations are specific to the outcome of interest. We could imagine that people in poorer health simply take more medications; many indicators of poor health, such as sedentary lifestyle, increase overall cancer risk. But, we expect that these people with poor health will also have increased risk of other non-cancer diseases. Therefore, the association would not be specific to cancer but would correspond to general sickliness. We create a set of negative control outcomes, matching each common cancer with a non-cancer outcome with a roughly similar incidence rate in our study population (*Figure 4A*). These outcomes are chosen to span a wide range of body systems. We apply the same hierarchical model to our negative control outcomes, which allows us to identify drug combinations that are generally associated with illness.

**Figure 4.**
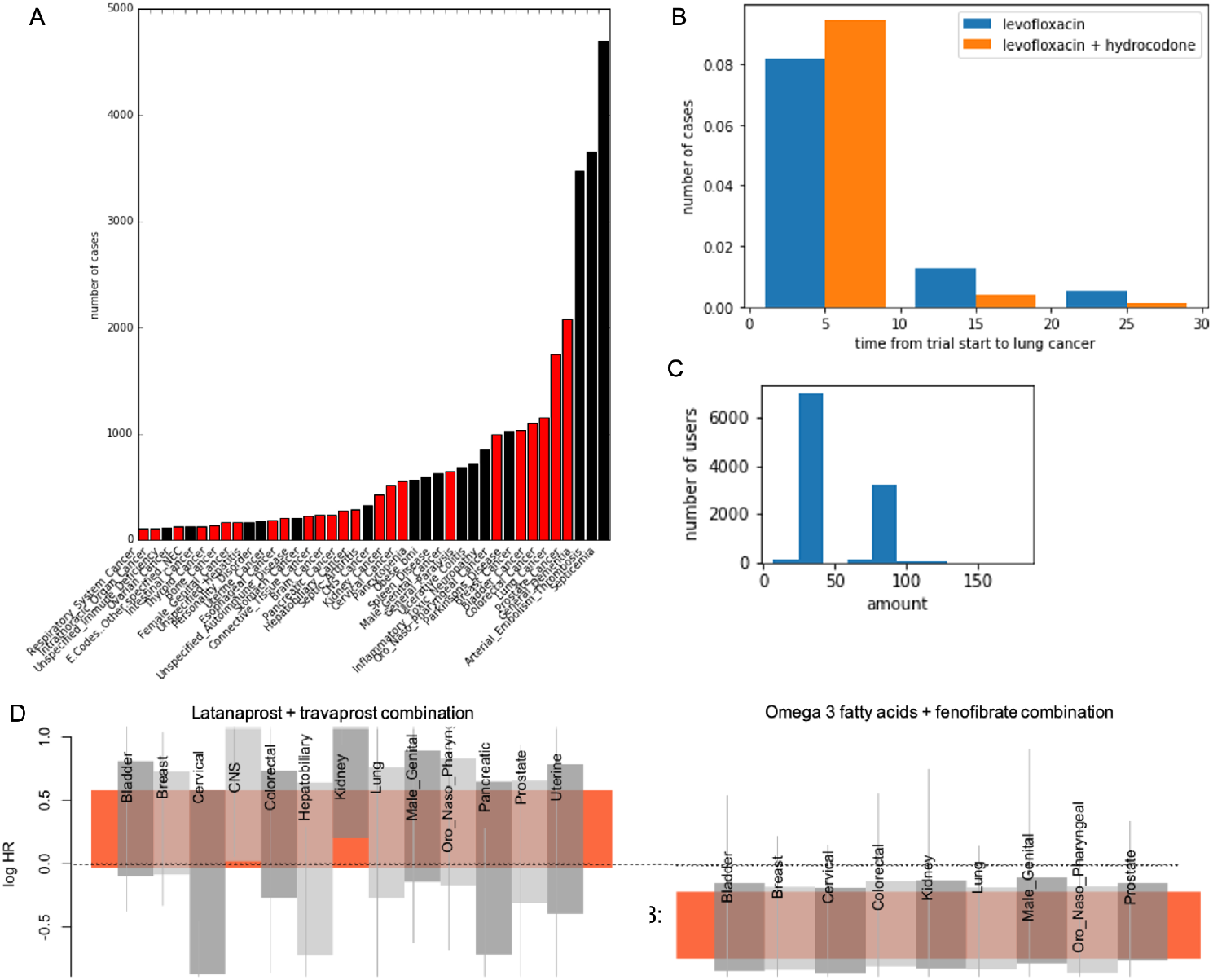
**A**. Overview of frequency of incidence of cancers (red) and of negative controls (black) (among one drug combination cohort study. **B**. Reverse causal effect of lung cancer on hydrocodone can be detected due to shorter time span from combination to cancer diagnosis (p = 8×10^−6^, rank sum test). **C**. Frequency of prescription for various amounts of fenofibrate. **D,E**. Integration of drug effect estimates (vertical lines, 95% Cl of log HR) in a hierarchical model: cohort study effects (vertical lines) are modeled aas samaples from effect per cancer type (gray boxes), which is in tern centered on aall-cancer effect (orange box, y-axis 95% Cl).

#### Detecting reverse causality

We find many examples of reverse causality, similar to the one illustrated in *Figure 3D*. As mentioned, we expect that these are due to erroneous missing information about a cancer diagnosis, where eventually the cancer diagnosis is recorded in the coded data, but only after some amount of treatment. It is reasonable to assume that when cancer in fact causes the treatment, the diagnosis of cancer will occur close in time to the treatment. We develop two tests to detect these cases. First, we test among cancer cases, whether the duration of time between the trial start date and date of cancer diagnosis is shorter among those treated with the combination than those in the comparator group (*Figure 4B*). Second, we test whether this temporal relationship is stronger for cancer types more strongly associated with the drug combination; that is, if drug combinations appear to have a stronger effect on cancers that also tend to occur more shortly after the combination is dispensed. In this manner, we can exclude the obvious cases of reverse causality.

### Sensitivity analyses to strengthen findings

#### Definition of exposure

After the previous series of tests, we are left with only a few significant associations from our drug-combination wide scan. We further subject these signals to a set of sensitivity analysis. These test whether the results are sensitive to some of the design decisions we made. We re-run the same drug wide association study for the candidate signals, twice, each with two different sets of assumptions. For each analysis, we perform the consistency analysis sketched in *Figure 3D* to assess whether the drug pair impacts all-cancer risk.

First, we change the window width used for defining trial start periods. While our primary design used 6 month windows, we create a secondary analysis that uses 3 month windows to define trial start periods. Note that this creates more follow-up periods, and results in a larger person-time matrix. Our second sensitivity analysis tests the effect of varying the definition of drug combination. While our main analysis censored person-time when the person stopped taking drug A, this results in a very limited follow-up time, where most people are followed for no more than a year. This can only allow us to detect short term drug effects. Removing this requirement, we can follow time after exposure for an average of 3.25 years, which is as long as people are followed for cancer outcomes in some randomized trials^5^. Removing the censoring greatly increases the number of cancers we can observe in follow up, but it also increases the size of our expanded person-time matrix. Both sensitivity analyses necessitate reduced numbers of people included in replicate cohort studies.

#### Amount of drug

Our final sensitivity analysis assesses the association of amount of drug B dispensed with cancer incidence. We find empirically that most drugs have a few common amounts dispensed, and we simply categorize users as in the “low” amount group or the “high” group. For instance, for fenofibrate users two main amounts dominate (*Figure 4C*). Because the number of people exposed to a drug combination and later getting cancer is rather low (*Figure 1D, Figure 3A,B*), we do not further sub-categorize drug amounts. We consider the signal has passed this test if 1) both categories are significant and 2) the effect size is stronger for the high-amount group than for the low group.

Combining the sensitivity analyses with the tests for artifacts, we narrow down our signals to the final associations. Our final results are presented in Supplementary Table 1 and Table 2 (see end). We find 50 single drug pairs are associated with either an elevated or a reduced rate of cancer. Combinations that do not have a consistent effect across most cancers can be excluded (*Figure 4D*). Among the combinations remaining, many can be further excluded from a cancer effect by the negative control test. Further, by combining signals across drugs in ingredient categories, we can prioritize drug combinations with a consistent effect in the ingredient class. Among our protective findings, only the combination of omega-3 fatty acids plus fenofibrate (*Figure 4E*) was able to be tested as an ingredient class (where fenofibrate is an ingredient of two drugs: fenofibric acid and fenofibrate). The ingredient class also was associated with reduced risk of cancer. As well, the sensitivity analysis showed that a higher amount of medication dispensed as associated with stronger protective effects as compared to the lower amount (average log hazard ratio of -2.9 for the high amount, versus -1.6 for the low amount).

## Discussion

We have described a systematic method to perform a drug combination-wide association study across all cancer types. While further studies are needed, one interesting finding is that a combination of Omega 3 acids (fish oil) with fenofibrate may be protective of cancer. Our negative control analysis indicates that this is not merely an effect of increased health consciousness but is specific to cancers. Our analysis of amount dispensed and other sensitivity analysis further supports the results over a 3 year follow up period. This pair of drugs both impact circulating lipid levels^20,21^. Changes to circulating lipids are known to influence cancer development, due to the high metabolic demands of growing tumors^22^. Therefore a biomedically relevant effect is plausible.

Our analysis has a number of limitations. Foremost, our results display evidence of remaining confounding. We previously showed that the propensity score is not able to model many important types of confounding^23^. This is because the medical record is incomplete. Many unpredictable factors can influence drug prescription, ranging from various comorbidities in medical history, to the patient’s gender, ethnicity, and geographical location^24^. Other studies have removed some sources of confounding by using methods that only compare each person to themselves across time^25,26^, but these are best suited to identifying acute short-term drug effects^27^. Therefore, future work must improve upon the propensity score and enable better modeling of confounding. Our results can provide a starting point for these efforts by providing a large set of cases of real-world confounding. A second limitation of our work is the limited follow-up time available for observing drug combination effects. In our analysis with no censoring, we have 3.25 years of follow-up time available. This is short, but it is in line with some previously combination drug effects discovered in randomized trials^5^. A third limitation is the small number of treated cases in some combinations of drugs. However, we are using one of the largest available sets of health claims data: therefore this issue would limit any study aiming to discover interactions from observational data. Finally, we use data covering the years 2003 to 2015, in order to avoid the change in the coding system to ICD-10. This should not impact our results, as any biological effect of a drug would not depend on year of administration, and all studies are limited to some time span.

Our unique approach has a number of strengths. First, this work represents, to our knowledge, the first application of marginal structural models for discovery of drug combinations impacting health outcomes. Marginal structural models are an established method to describe effects of time-varying exposures on health outcomes. But, our unique design implements these methods to describe the time-varying effect of adding a second drug on top of treatment with an initial drug. Another unique method is our hierarchical model for combining estimates of drug effects on related diseases to create an overall summary of the effect on a disease category. While some other studies have combined drug effect estimates using a hierarchical model^28^, our study is the first to use hierarchical models to combine effects on related diseases to create an effect estimate robust to some types of confounding.

A final and crucial strength of our design is its systematic nature. By performing a comprehensive analysis across all testable drug combinations, we are able to compare methods and discover their weaknesses. For instance, we showed that methods that only account for a few general confounders are quite vulnerable to bias. Our thorough consideration of sensitivity to study design, and our attempts to quantify remaining confounding, increase confidence in our results. We expect that this approach can provide a basis for future work seeking to discover drug combination effects from health data.

## Methods

### Expansion of medical record into person time

In order to account for time-varying confounding, we follow the approach of Danaei, et. al in which participant time after enrollment (initiation of drug A) is used to form a sequence of randomized trials^15,16^. We divide the health record into windows of time since initiation of drug A. Our main analysis uses windows of 24 weeks, and a subsequent sensitivity analysis uses windows of 12 weeks. For each window *t*, we obtain time-specific data for patient *p*, denoted *health*_*p,t*_, including if they started drug B (*treatment*_*p,t*_), and other medical information at that time, including incidence of any cancer. As outlined in *Figure 2*, we expand person-time into repeated trials. Specifically, people are eligible for enrollment into a trial at window *t* as long as 1) they are still observed in the data set (and not censored, depending on the design) and 2) they have not yet taken drug B, and they have not yet had the outcome of interest (here, a specific cancer). In trial 1, the person’s full time observed is the amount of time available for follow-up: all time windows are included. For subsequent trials, follow-up time begins only after trial initiation. In Table 1, we illustrate the expansion into person-time for person i and person v from *Figure 2*. Therefore, the time intervals are used for two purposes: 1) to assess whether a person remains eligible for a trial, and which treatment arm they fall into in the trial; and 2) to follow the outcomes of people after enrollment into a trial. The shorter the windows are, the more rows this table will have. The longer the follow-up time, the more rows this table will have. Eligibility time and follow-up time increase when no censoring is performed, resulting in a larger person-time table.

**Table 1:** Expansion into trials

| Table 1: Expansion into trials |  |  |  |  |  |  |  |
| --- | --- | --- | --- | --- | --- | --- | --- |
| Person | Trial | Follow-up | Treatment | Outcome | Treatment weight model | Treatment probability | comment |
| i | 1 | 1 | A | no | yes | $p(A+B_{i,1}=False)$ | person i is not eligible for trials after trial 2 because they took the combination at trial 2 |
| i | 1 | 2 | A | no | no | $p(A+B_{i,1}=False)$ | |
| i | 1 | 3 | A | no | no | $p(A+B_{i,1}=False)$ | |
| i | 1 | 4 | A | yes | no | $p(A+B_{i,1}=False)$ | |
| i | 2 | 1 | A+B | no | yes | $p(A+B_{i,1}=False) \times p(A+B_{i,2}=True)$ | |
| i | 2 | 2 | A+B | no | no | $p(A+B_{i,1}=False) \times p(A+B_{i,2}=True)$ | |
| i | 2 | 3 | A+B | yes | no | $p(A+B_{i,1}=False) \times p(A+B_{i,2}=True)$ | |
| v | 1 | 1 | A | no | yes | $p(A+B_{v,1}=False)$ | person v is only followed for 2 windows, they have no drug B or outcome in either. |
| v | 1 | 2 | A | no | no | $p(A+B_{v,1}=False)$ | |
| v | 2 | 1 | A | no | yes | $p(A+B_{v,1}=False) \times p(A+B_{v,2}=False)$ | |

### Modeling confounding

We follow the marginal structural model approach, which adjusts for confounding by modeling both time-varying confounding and baseline confounding of treatment. For each person *p*, for each trial *t*, we estimate probability of the person participating in the drug combination arm, rather than the single-drug arm, given medical history: *p*(*treatment*_*p,t*_|*health*_*p,t*_, b*aseline info*_*p*_)

Then, in the marginal structural model, we use these probabilities to create treatment weights. The same weights apply to the entire trial.

First, we create the model to estimate the probability of treatment given health history. All person-time windows eligible for enrollment are included in the model, but not the expanded follow-up time of each person: in Table 3, all rows where “Treatment weight model” is “yes” are included in model fitting. The outcome variable in the logistic regression would be the contents of the “Treatment” column in that table. Typically, these models are built with a few selected confounders. But, because we do not wish to assume that these are the only confounders, here, we implement this model with a sparse high-dimensional logistic regression. We include all treatments and diagnoses as variables in *health*_*t*_ and *baseline*. Baseline info also includes gender and cubic spline variables to model the effect of age, calendar time, and the number of total diagnoses and treatments the person has, as a measure of overall medical burden.

Typically, our data has over 10,000 features, but we remove the features observed in fewer than 100 people. We fit the model using elastic net logistic regression using the scikit learn package, performing a grid search with cross-validation to choose values for the regularization parameter and the l1-ratio. For each model, we perform the hyperparameter tuning separately. After regularization, typically only a hundred or fewer features have a non-zero coefficient in the logistic regression model. Finally, we obtain the predicted probabilities of treatment from the model. Then, we create a marginal structural model to estimate the association of treatment with a cancer outcome. In this method, each person-trial is weighted based on the probability of initiating treatment at that trial (as calculated above), and no earlier, given their medical history. So, for person *i* who initiated the drug combination at trial 2, they had to first not initiate the combination in time 1, then initiate in time 2, so the total probability of initiating treatment at time 2 is *p*(*A*+ B_*i*,1_ =*False*)×*p*(*A*+ B_*i*,2_ =*True*). Therefore, to calculate the probability of the sequence of history that resulted in that person’s enrollment in to the second trial, we multiply the appropriate time window probabilities to obtain the overall probability:

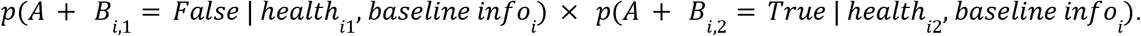

Finally, we transform the probabilities to weights: the inverse of the resulting probabilities are used as *treatment weights* for each trial. Since previous publications in marginal structural modeling proved that using the *stabilized weights* reduces extreme weights without adding bias, we use stabilized weights. All follow-up time windows in the trial receive this same treatment weight (see Table 3).

### Censoring

In our primary analysis, we censor people when they discontinue drug A. This is a design choice that allows us to estimate the effect specifically of simultaneous exposure to drugs A and B, rather than including people with possibly sequential exposure. (Other designs are tested in the sensitivity analyses). Time-varying confounding could cause someone to stop taking the drug, influencing censoring and potentially biasing our results^8,29^. To account for time-varying confounding of censoring, as with the treatment weights, we again model the relationship between medical history and censoring, and use this to weight each person-time observation that could be censored. Again we follow the method outlined in^8^, where we weight each person-time observation by modeling probability of censoring given medical history. If someone is *not* censored, they did *not* discontinue drug A, so we model *p*(*discontinue drug A*_*p,t*_ =*False*|*health*_*p,t*_, b*aseline info*_*p*_) using a high-dimensional sparse logistic regression model, as in the creation of treatment weights. Finally, we obtain the weights in the same manner described above. All person-time follow-up is eligible for censoring (and censoring weights are the same for the same person-time window that appears in different trials). To weight each observation adjusting for confounding of discontinuation of drug A and of treatment with drug B, we multiply the two weights together. While this makes the assumption that the two treatment decisions are independent, by performing the sensitivity analysis without censoring, we are able to assess whether that assumption impacts the results.

### Estimating time to cancer in survival analysis

Once we obtain our weights, they can be applied in the weighted Cox regression using the R *survival* package^8,30^. Specifically, our implementation uses the time interval encoding of time-to-event data to indicate a subject’s presence in the risk set and their treatment and outcome (incidence of a cancer type) at each interval. We apply weights to each interval. This part of our analysis follows the approach previously implemented in other studies modeling a sequence of randomized trials with observational data^15,16,31^. In order to account for the repeated presence of subjects in multiple trials, we use the “cluster” option, treating each subject as one clustered set of observations^32^. Finally, we obtain hazard ratios and robust standard errors, using the R survey package.

### Hierarchical model

The hierarchical model shown in *Figure 3E* is presented in Appendix Model. The slightly simpler version without the ingredient layer is very similar, as it only removes this layer of the hierarchy. The model uses as input the log hazard ratios of the treatment effects estimated from the Cox regression model. These are assumed to have a normal distribution in the Wald test. Our model posits that these estimates are samples derived from a normal distribution centered around a true effect estimate. We fit the model using Stan, and obtain the posterior distribution of the all-cancer effect from this model. We use the same model to obtain the effect estimate across all negative control outcomes, replacing the cancer estimates with the estimates of the effect of each drug combination on incidence of each negative control health condition.

### Sensitivity analyses and other tests

We repeat our drug-wide analysis, but only testing drugs that have effects tested in our main analysis. For the first sensitivity analysis, we alter the size of the windows, and for the second sensitivity analysis, we do not censor when drug A is discontinued. Because, as mentioned above, both of these alterations can result in large increases in the size of the expanded person-time data set, we must reduce the number of drug A users included in some analyses. Therefore, we split the initial cohort into replicate cohort studies of independent cohorts; the results are pooled in the hierarchical model.

To perform the reverse causality test, we gather data on time to cancer for the cancers most strongly associated with the drug combination. Considering only the people who eventually get that type of cancer, we use the rank-sum test to evaluate whether the time to cancer is significantly shorter for those taking the combination versus those in the single-drug group.

These values are presented in Table 1 and 2, “reverse causality test” column. To categorize combination users into those with “low” and “high” amount of the second drug, we obtain the two most common amounts prescribed. We set the cut point to be half way between those two amounts. Then, we encode all people as non-users, low users, and high users, and obtain the effect of low or high drug use.

## Data Availability

All data produced in the present study are available upon reasonable request to the authors

## Code availability

Python code to form the sequence of repeated trials, and R code to perform the survival analysis will be provided alongside the code to run our stan model, at https://github.com/RDMelamed/drug-combo-cancer.git

**Supplementary Table 1:**
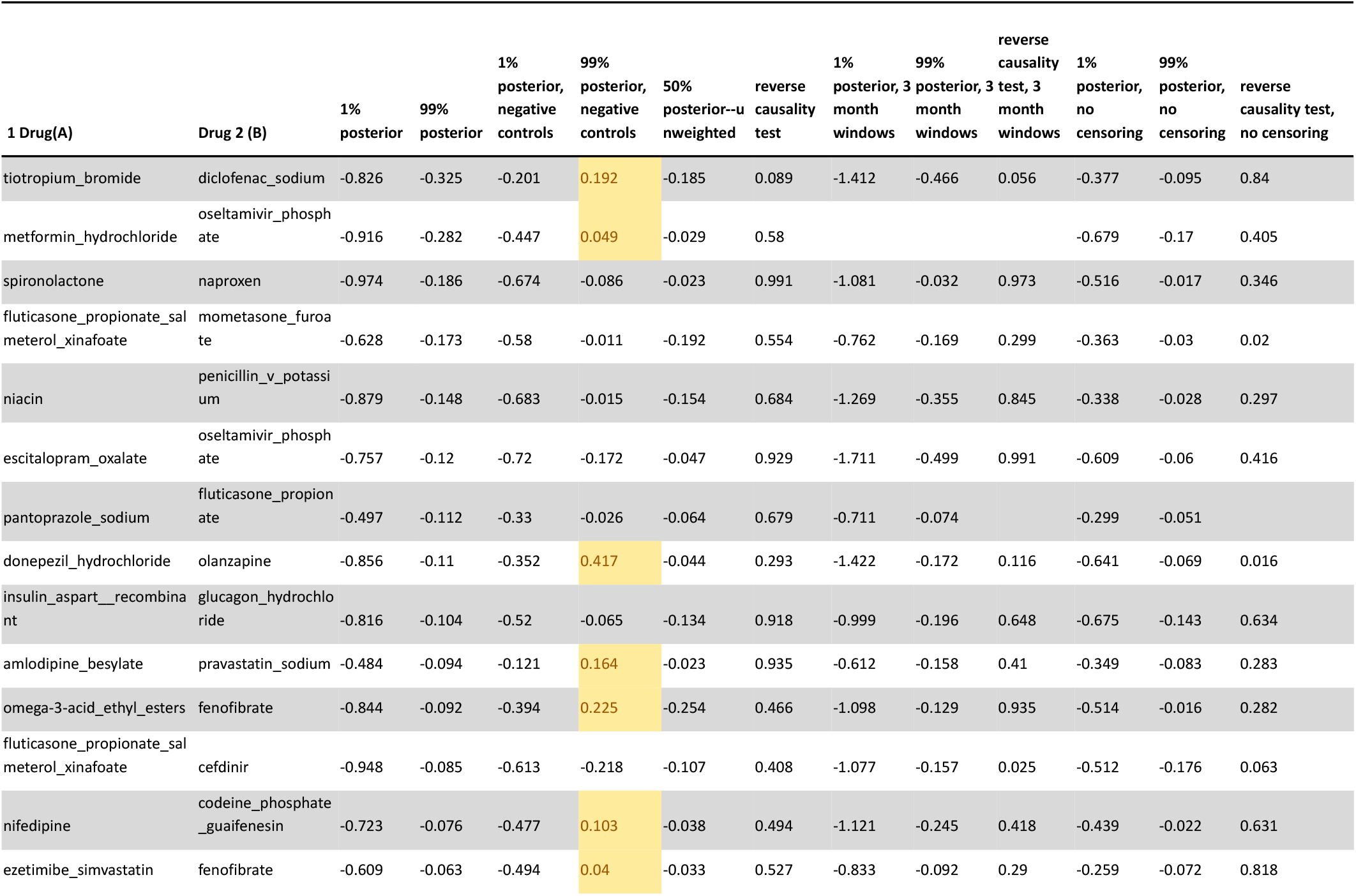
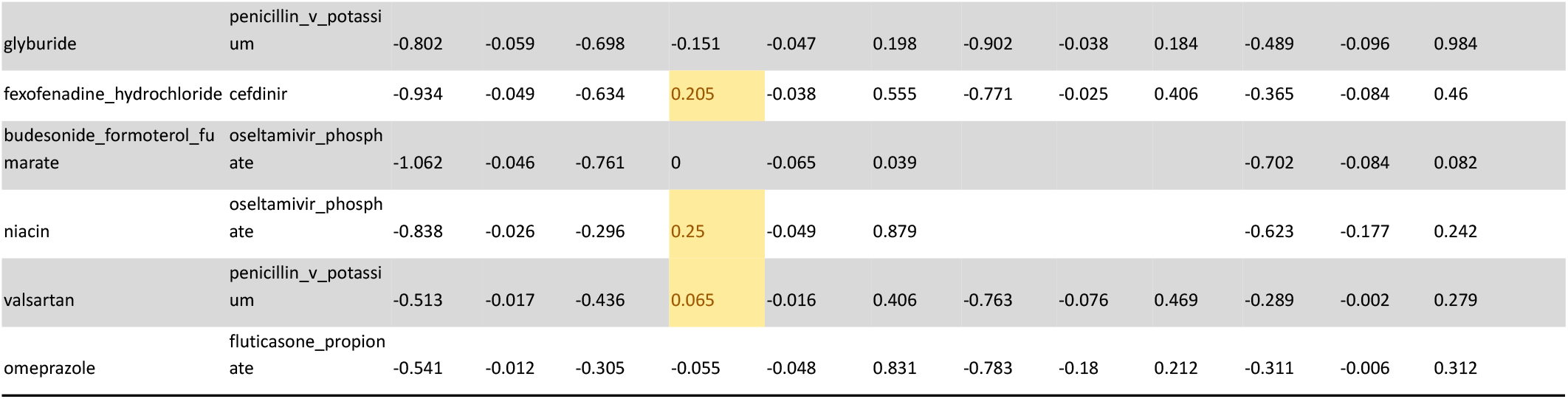
Drug combinations associated with reduced risk of cancer. The negative control test is passed for those in yellow (no reduced risk of non-cancers)

**Supplementary Table 2:**
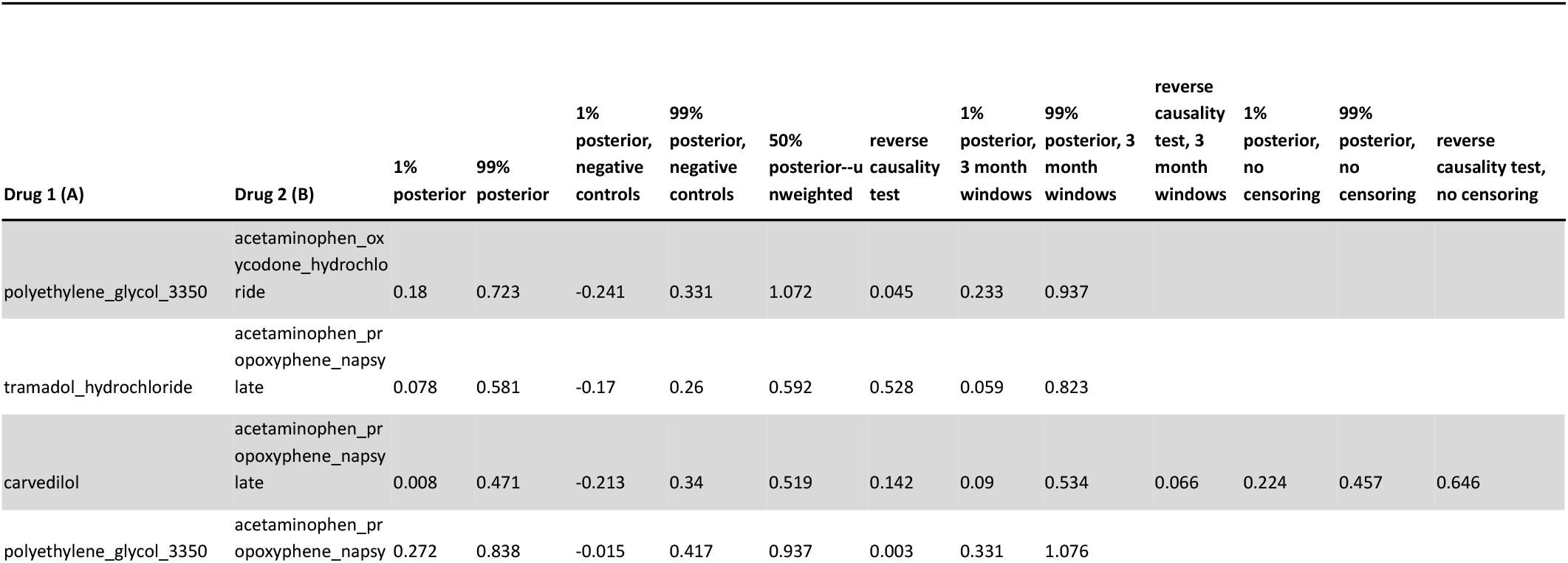
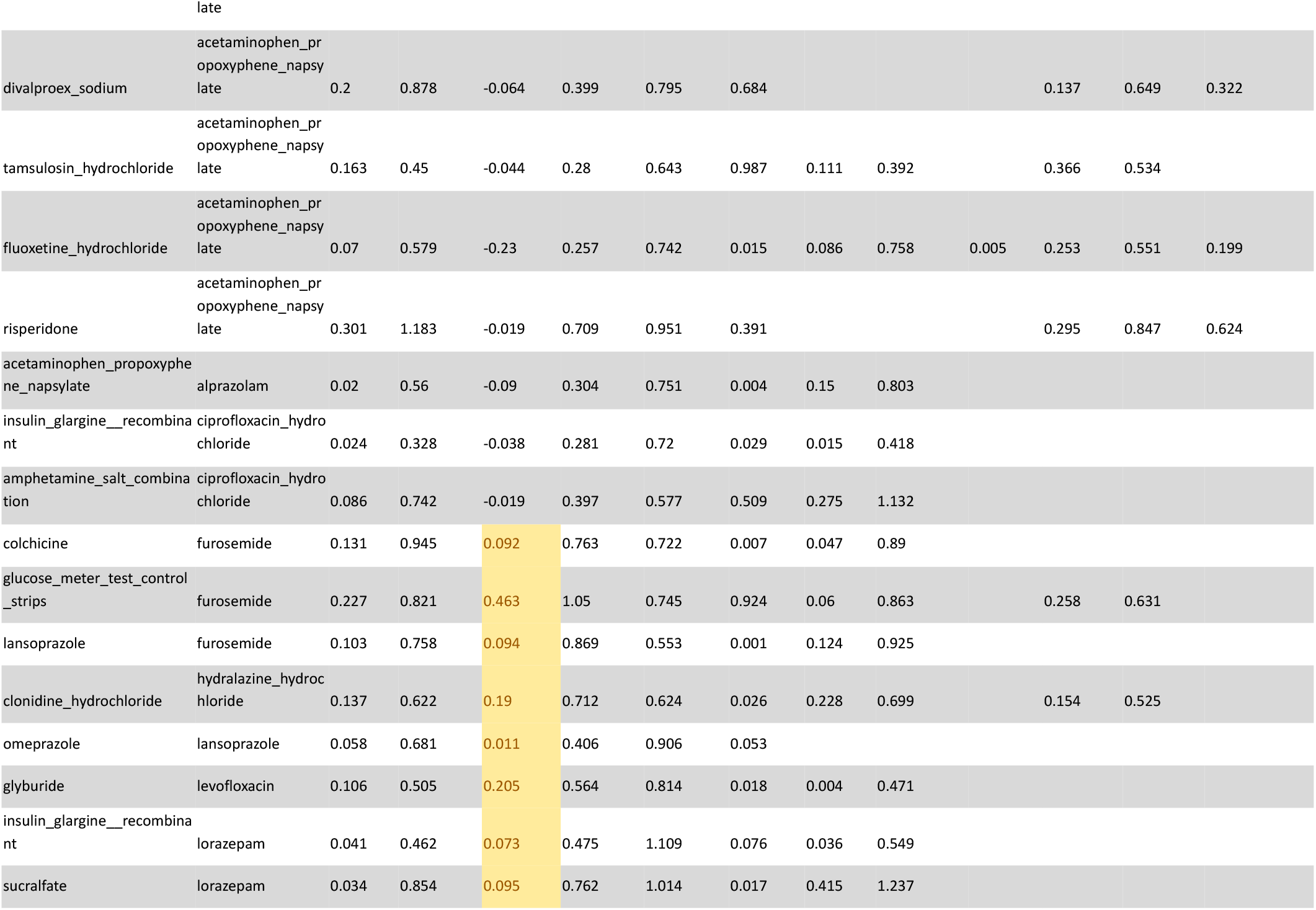
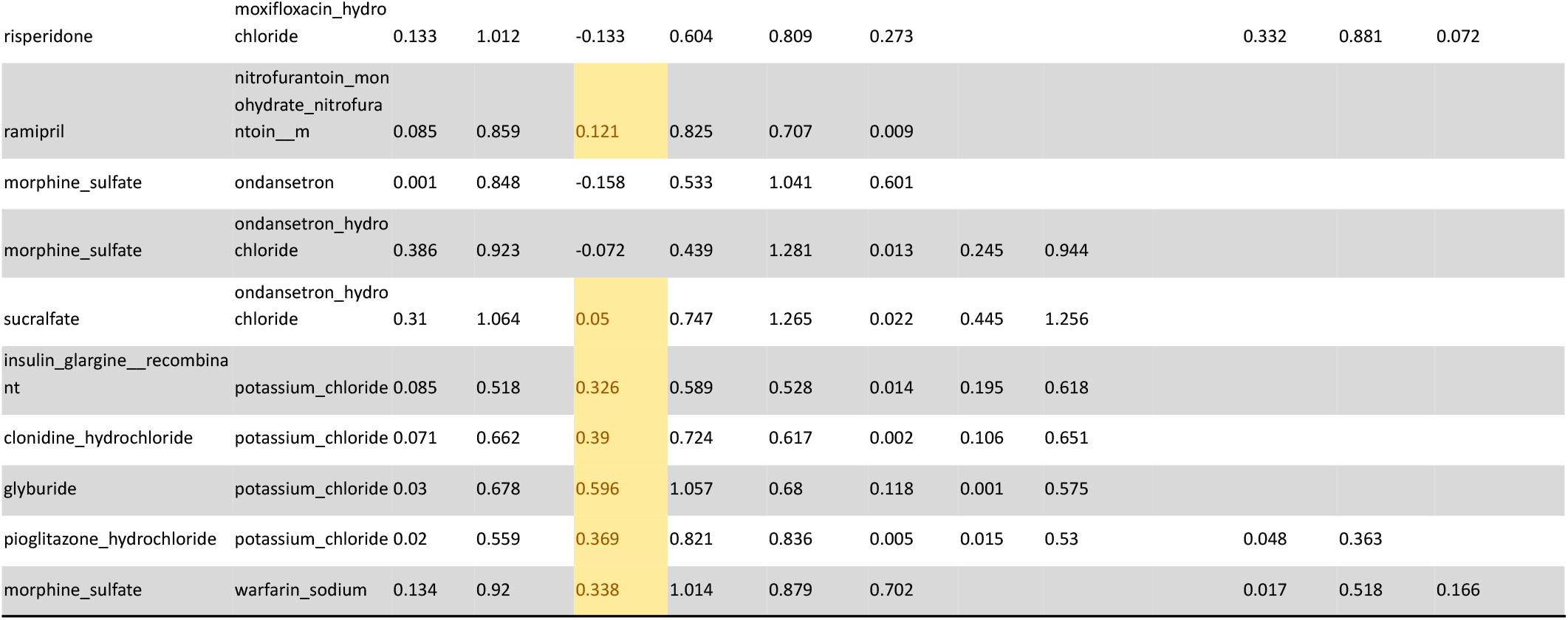
Drugs combinations associated with increased risk of cancer. The negative control test passes for those in yellow (no increased risk of non-cancer). But, most of these are cancer therapies and likely due to reverse causality.

## Supplementary Model

~~~
data{
  ///
  int <lower=1> N; // tot
  int <lower=1> Noutcome;
  real cancer_effect_sd; // prior variance for mean of eff
  real percancer_devsd; // prior variance for mean of eff
  real across_cancers_sd; // prior variance for mean of eff
  vector[N] expvals;
  vector[N] se;
  //int<lower=0, upper=1> outcome_is_cancer[Noutcome];
  int N_class;
  int<lower=1,upper=N_class> drug2[N];
  //int<lower=0, upper=1> cancer_indicator[N];
  int<lower=1, upper=Noutcome> outcome_id[N];
  }
  parameters{
  vector[N_class] eff_per_drug_cancer_tilde[Noutcome];
  real class_cancer_effect;
  real<lower=0> class_allcancer_sd;
  vector<lower=0>[Noutcome] outcome_sd_across_drugs; // for each outcome, the spread across drugs might be different
  vector[Noutcome] class_per_cancer_effect_tilde;
  }
  transformed parameters{
  real Xeff_rep[N];
  vector[N_class] eff_percancer[Noutcome];
  vector[Noutcome] class_per_cancer_effect = class_per_cancer_effect_tilde * class_allcancer_sd
+ class_cancer_effect;
    for(i in 1:Noutcome){
     eff_percancer[i] = eff_per_drug_cancer_tilde[i] * outcome_sd_across_drugs[i] +
  class_per_cancer_effect[i];
  }
  for (i in 1:N){
        Xeff_rep[i] = eff_percancer[outcome_id[i]][drug2[i]] ;
    }
  }
  model {
  // these 2 are the effect & var of effect overall of cancer of drugs in that group
  class_cancer_effect ∼ normal(0, cancer_effect_sd);
  class_allcancer_sd ∼ normal(0,across_cancers_sd); // overall, how wide is the spread across cancers
  // then these are the variations across cancers of that
  class_per_cancer_effect_tilde ∼ normal(0, 1);
  // then we get to the per-drug variation from that for each cancer: the random deviations,
  and what we scale these by per cancer
  outcome_sd_across_drugs ∼ normal(0, percancer_devsd);
  for(i in 1:Noutcome){
        eff_per_drug_cancer_tilde[i] ∼ normal(0,1); // each of these being random spread around
the per-cancer effect, across drugs in that class
  }
     if(run_estimation==1){
      expvals ∼ normal(Xeff_rep, se); }
  }
~~~

## Notes

*Additional information:* This work was supported by K01ES028055 the National Institutes of Health to RDM.

The authors declare no conflicts.

### Competing Interest Statement

The authors have declared no competing interest.

### Funding Statement

K01ES028055

### Author Declarations

This study used only deidentified publicly available human claims data from MarketScan IBM and has been deemed to be not human subjects research.

### Summary of Updates

Text has been edited for clarity and figures merged for ease of viewing.

